# Characterizing COVID-19 case detection utilizing influenza surveillance data in the United States, January-March, 2020

**DOI:** 10.1101/2020.04.23.20077651

**Authors:** Micaela Sandoval, Adam Hair, Shreela Sharma, Catherine Troisi

## Abstract

COVID-19 reached the US in January, 2020, but state and local case detection efforts varied in timing and scale. We conducted a state-level ecological analysis of COVID-19 epidemiology alongside CDC influenza surveillance data and policy timelines. Our findings show wide variation in COVID-19 case detection and influenza-like-illness activity between states.

## Introduction

The COVID-19 pandemic, which originated in China in late 2019, has since emerged as the dominant public health threat facing the global community. Early data from China and Europe suggested a case fatality rate of 1-3% and a hospitalization rate of 9-11%, but these measures are derived from highly variable case estimates [1, 2]. COVID-19 control and prevention has been further complicated by an extended incubation period and proportion of asymptomatic transmitters, which impedes real-time surveillance and response efforts[3, 4]. Data from the contained Diamond Princess cruise outbreak showed an estimated asymptomatic proportion of 15-20%, and data from Singapore suggests significant transmission by pre-symptomatic cases[5, 6]. As essential epidemic variables remain unknown, public health authorities in many countries have focused on case detection, contact tracing, and isolation measures to reduce morbidity and mortality due to COVID-19[7, 8].

The United States reported its first COVID-19 case in late January, 2020; as of April 20, 2020, the US has confirmed 786,968 cases of COVID-19[9]. However, in the absence of population-based surveillance data, case estimates depend on individual state and local health departments’ testing strategy and sufficiency of testing. Therefore, public health researchers must draw on established surveillance systems to estimate the true disease burden; investigators in Europe utilized influenza surveillance to monitor excess influenza-like illness during the outbreak[10]. We conducted a state-level ecological analysis in four US states with high COVID-19 incidence: Washington, Colorado, Louisiana, and New York, to examine the relationship between influenza-like illness and COVID-19 morbidity and mortality. This method may be a valuable way to estimate prevalence of a new ILI in a population early in an outbreak when testing capacity is limited.

## Methods

We conducted a state-level ecological analysis of influenza like illness, pneumonia mortality, and COVID-19 epidemiology. We focused on US states with high COVID19 incidence representing variable testing strategies to examine the temporal relationship between trends in respiratory surveillance data and the COVID-19 outbreak. COVID-19 confirmed case counts, death counts, and testing data were retrieved from the Johns Hopkins University & Medicine Coronavirus Dashboard on April 16, 2020[9].. Cumulative incidence and testing rates were calculated using national and state population estimates for January 1, 2020[11]. Policy data was retrieved from the Boston University School of Public Health State policy tracker dataset on April 18, 2020[12].

Influenza surveillance data were retrieved from the Centers for Disease Control and Prevention FluView dashboard on April 17, 2020[13]. Influenza testing data is collected from representative clinical and public health laboratories participating in the U.S. WHO Collaborating Laboratories System and the National Respiratory and Enteric Virus Surveillance System (NREVSS). Only clinical laboratory surveillance data were used for this analysis as they are more representative of symptomatic patients presenting for care. Pneumonia and influenza mortality data is collected from state vital statistics offices by the National Center for Health Statistics using ICD-10 multiple cause of death codes[13]. This analysis included raw pneumonia death counts to reduce bias resulting from the inclusion of confirmed influenza deaths. Comparisons between geographic areas utilizing unweighted surveillance data is not appropriate, so this investigation focuses on characterizing each state’s population individually. CDC influenza surveillance data was deemed unreliable by investigators after MMWR week 12 (March 22, 2020), and at time of writing, FluView has been abbreviated for the remaining season[13].

## RESULTS

Our analysis focused on four US states with high COVID-19 incidence. Cumulative incidence (January 22 to April 16, 2020) varied widely from 144 cases to 1,150 cases per 100,000 population, despite homogeneity in stay at home order dates (Table 1). Of the selected states, only Washington exhibited a higher than average number of negative influenza tests resulted by clinical laboratories participating in the WHO or NREVSS systems in the first eight weeks of 2020 (Figure 1). In Washington, New York, and Louisiana, week 12 increases in reported negative influenza tests were eclipsed by incident COVID-19 cases. However, in Colorado, negative influenza tests increased markedly from week 9 to week 12.

**Table 1.** COVID-19 Epidemiology and Policy Timeline, selected US states

|  | Incidence<br>per 100k | Case<br>fatality | Tests per<br>100k | # tests/<br>case* | Hospitalization<br>rate | State of<br>emergency date | Stay home<br>order date |
| --- | --- | --- | --- | --- | --- | --- | --- |
| Colorado | 144 | 4.3% | 704 | 4.9 | 20% | 3/11/2020 | 3/26/2020 |
| Washington | 145 | 5.2% | 1,632 | 11.2 | 5.4% | 2/29/2020 | 3/23/2020 |
| Louisiana | 485 | 5.1% | 2,723 | 5.6 | 8.5% | 3/11/2020 | 3/23/2020 |
| New York | 1,150 | 6.6% | 2,830 | 2.5 | 23% | 3/7/2020 | 3/22/2020 |
| United States | 203 | 4.9% | 1,033 | 5.1 | 16.3% |  |  |

COVID-19 case data retrieved from Johns Hopkins University dashboard April 16, 2020. State policy data retrieved from Boston University policy dataset, April 18, 2020.
\*Number of tests needed to detect one case = (Total population tested / confirmed cases)

**Figure 1.**
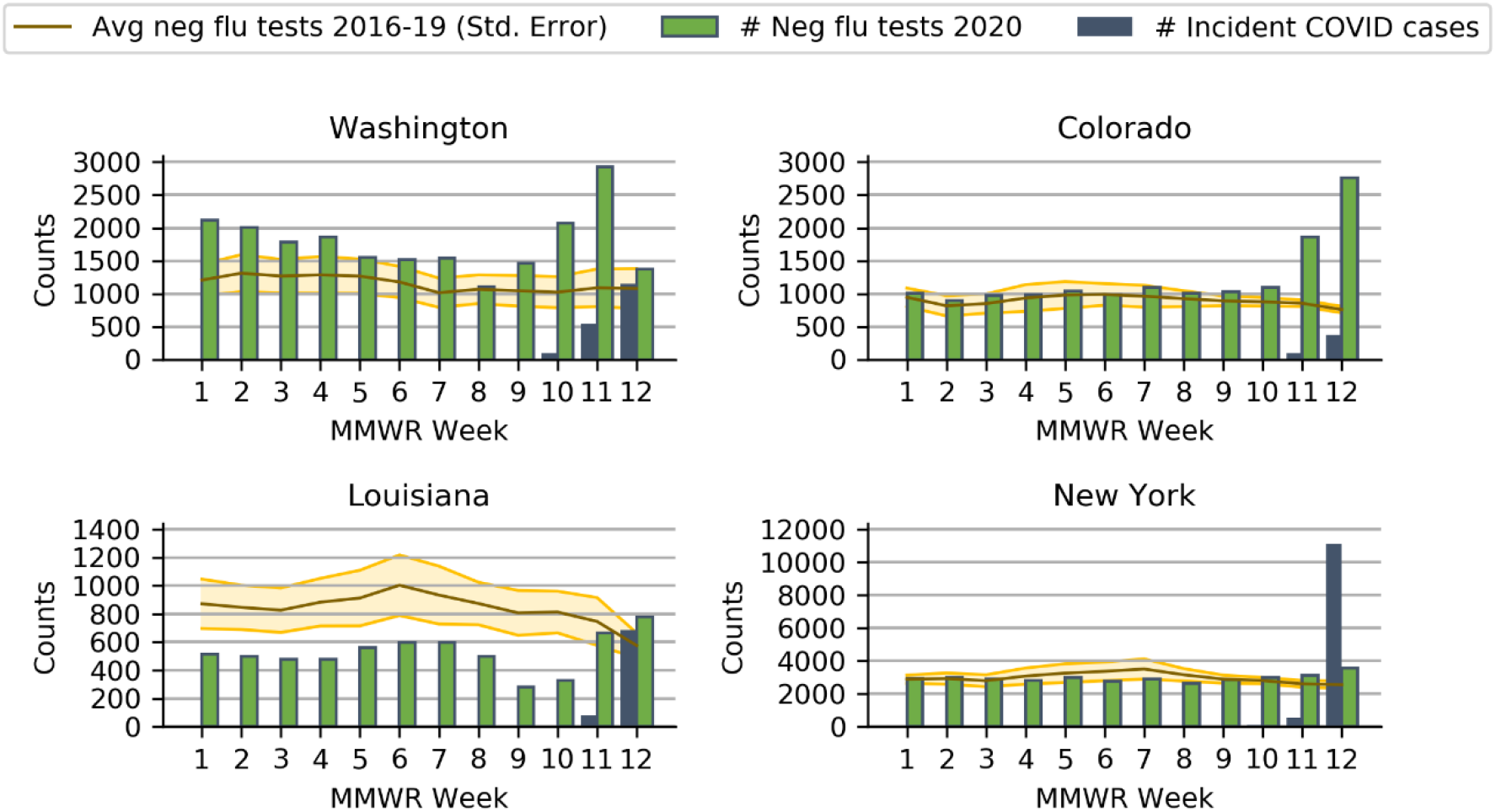
Influenza Surveillance and COVID-19 incidence: January-March, 2020. COVID-19 data retrieved from Johns Hopkins University dashboard, April 16, 2020. Influenza surveillance data retrieved from CDC Influenza Surveillance dashboard, April 17, 2020.

Testing volume in MMWR weeks 10-12 varied widely between states (Figure 2). Notably, in Colorado, testing volume was low in relation to negative influenza tests. With the exception of Washington, states did not document significant COVID-19 mortality until week 12 (Table 2). Colorado and Washington demonstrated higher than average pneumonia deaths in week 12.

**Table 2.** COVID-19 and Pneumonia Mortality, selected US states

|  | MMWR<br>Wk 9 | MMWR<br>Wk 10 | MMWR<br>Wk 11 | MMWR<br>Wk 12 |
| --- | --- | --- | --- | --- |
| **Colorado** |  |  |  |  |
| COVID-19 deaths | 0 | 0 | 1 | 4 |
| Pneum. deaths, 2020 | 53 | 46 | 35 | 65 |
| Mean pneum. deaths<br>2016-2019 | 44 | 56 | 53 | 49 |
| **Washington** |  |  |  |  |
| COVID-19 deaths | 1 | 26 | 24 | 57 |
| Pneumonia deaths 2020 | 73 | 96 | 85 | 121 |
| Mean pneum. deaths<br>2016-2019 | 75 | 84 | 73 | 88 |
| **Louisiana** |  |  |  |  |
| COVID-19 deaths | 0 | 0 | 0 | 16 |
| Pneumonia deaths 2020 | 39 | 31 | 38 | 39 |
| Mean pneum. deaths<br>2016-2019 | 50 | 48 | 50 | 47 |
| **New York** |  |  |  |  |
| COVID-19 deaths | 0 | 0 | 0 | 44 |
| Pneumonia deaths 2020 | 152 | 160 | 169 | 198 |
| Mean pneum. deaths<br>2016-2019 | 169 | 168 | 163 | 178 |

COVID-19 case, death, test, and hospitalization data retrieved from Johns Hopkins University dashboard, April 16, 2020.
Pneumonia mortality data retrieved from CDC Influenza Surveillance dashboard, April 17, 2020.

**Figure 2.**
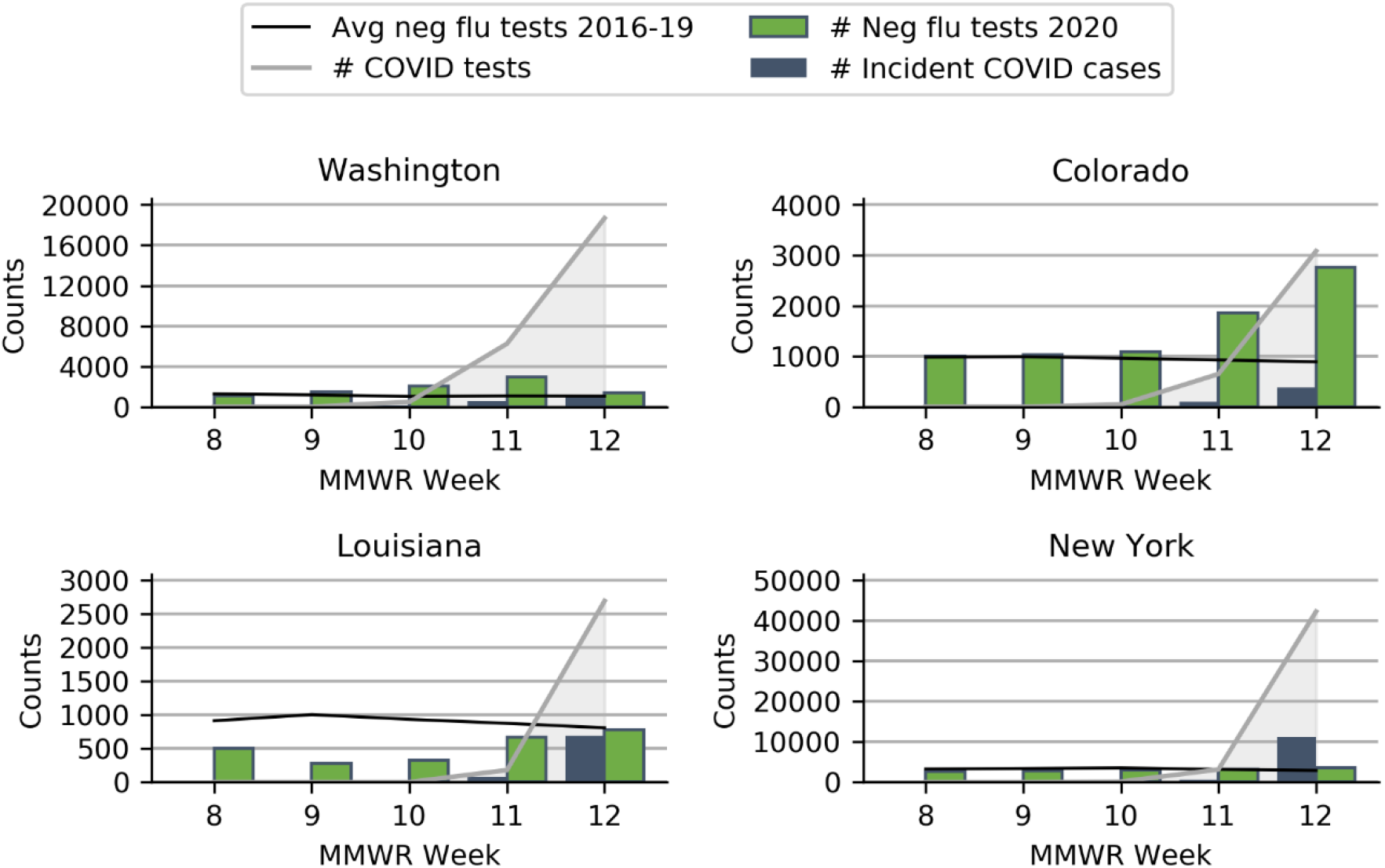
Influenza Surveillance, COVID-19 incidence, and COVID-19 testing: January-March, 2020. COVID-19 data retrieved from Johns Hopkins University dashboard, April 16, 2020. Influenza surveillance data retrieved from CDC Influenza Surveillance dashboard, April 17, 2020.

While cumulative incidence was similar in Colorado and Washington (144 and 145 cases per 100,000 pop, respectively), testing rates in Colorado were less than half testing rates in Washington, and hospitalization rates in Colorado were 3.7 times hospitalization rates in Washington. Additionally, Washington and Colorado both reported negative influenza tests in excess of the 2016-2019 mean for weeks 11 and 12, but confirmed COVID-19 cases accounted for almost 80% of excess negative influenza tests in Washington and only 16% of excess tests in Colorado. In weeks 10 to 12, Washington implemented COVID-19 testing at 334 tests per 100,000 population, while Colorado conducted only 66 tests per 100,000 population. In contrast, neither New York nor Louisiana exhibited significant excess negative influenza tests or pneumonia mortality during MMWR weeks 1 to 12. While both states utilized high testing rates, New York’s incidence and hospitalization rate were 2.4 and 2.7 times Louisiana’s, respectively. At the time of writing this manuscript, the New York City metropolitan area bears the highest disease burden in the US, and NYC caseloads drive the epidemiologic characteristics of the country.

## Discussion

This ecological analysis was conducted on publicly available surveillance data amidst a rapidly evolving outbreak. Confounding from differential surveillance reporting, prevalence of risk factors, differences in social distancing guidance, and changes in health-seeking behavior is likely. The data presented is not representative of the United States and should not be extrapolated. Although we were not able to estimate influenza-like-illness burden or pneumonia mortality throughout the outbreak, the results of this hypothesis-generating analysis provide compelling evidence for further studies.

The proportion of confirmed COVID-19 cases requiring hospitalization is impacted by both disease severity in the population and case detection rate. The extraordinarily high hospitalization rates and in Colorado and New York likely belie a large population of undetected cases. Case fatality rates may be influenced by health system capacity in addition to detection bias. While passive surveillance and COVID-19 testing data cannot be linked at an individual level due to the ecological nature of the study, sharp increases in negative influenza tests correspond to additional patients presenting with symptoms such as fever, cough, sore throat, and body aches for whom influenza is not present. Excess pneumonia mortality likewise suggests an increase in severe respiratory infections, presumably COVID-19. Colorado’s epidemiologic profile in particular demonstrates a concerning discrepancy between influenza like illness disease burden and reported incidence of COVID-19.

Our findings depict startling heterogeneity in COVID-19 surveillance as defined by testing across high-incidence states. The volume of negative influenza tests and pneumonia mortality data likely underestimate true rates of influenza-like-illness and mortality, due to lags in reporting, changing health-seeking behaviors, test shortages, and rising telehealth modalities. Additionally, these proxies estimate symptomatic cases; evidence shows COVID-19 transmission is largely precipitated by asymptomatic or pre-clinical carriers[14]. Our findings indicate a vast deficit in COVID-19 detection which must be resolved before transmission can be effectively contained.

## Data Availability

Data is publicly available from referenced sources

## REFERENCES

1. Li, Q., et al., Early Transmission Dynamics in Wuhan, China, of Novel Coronavirus–Infected Pneumonia. New England Journal of Medicine, 2020. 382(13): p. 1199–1207.

2. Remuzzi, A. and G. Remuzzi, COVID-19 and Italy: what next? The Lancet, 2020. 395(10231): p. 1225–1228.

3. Fauci, A.S., H.C. Lane, and R.R. Redfield, Covid-19 — Navigating the Uncharted. New England Journal of Medicine, 2020. 382(13): p. 1268–1269.

4. Lipsitch, M., D.L. Swerdlow, and L. Finelli, Defining the Epidemiology of Covid-19 — Studies Needed. New England Journal of Medicine, 2020. 382(13): p. 1194–1196.

5. Wei WE, L.Z., Chiew CJ, Yong SE, Toh MP, Lee VJ, Presymptomatic Transmission of SARS-CoV-2 — Singapore, January 23–March 16, 2020. MMWR Morb Mortal Wkly Rep, 2020. 69:: p. 411–415.

6. Mizumoto, K., et al., Estimating the asymptomatic proportion of coronavirus disease 2019 (COVID-19) cases on board the Diamond Princess cruise ship, Yokohama, Japan, 2020. Eurosurveillance, 2020. 25(10): p. 2000180.

7. Hellewell, J., et al., Feasibility of controlling COVID-19 outbreaks by isolation of cases and contacts. The Lancet Global Health, 2020. 8(4): p. e488–e496.

8. Ng, Y., et al., Evaluation of the Effectiveness of Surveillance and Containment Measures for the First 100 Patients with COVID-19 in Singapore - January 2-February 29, 2020. MMWR Morb Mortal Wkly Rep, 2020. 69(11): p. 307–311.

9. Dong, E., H. Du, and L. Gardner, An interactive web-based dashboard to track COVID-19 in real time. The Lancet Infectious Diseases.

10. Boelle, P.Y., et al., Excess cases of influenza-like illnesses synchronous with coronavirus disease (COVID-19) epidemic, France, March 2020. Euro Surveill, 2020. 25(14).

11. 2019 National and State Population Estimates. 2019, United States Census Bureau: Washington, DC.

12. Raifman, J., et al., “COVID-19 US state policy database”. 2020, Boston University School of Public Health: Available at: www.tinyurl.com/statepolicies.

13. Overview of Influenza Surveillance in the United States. 2020, Centers for Disease Control and Prevention: Atlanta.

14. Bai, Y., et al., Presumed Asymptomatic Carrier Transmission of COVID-19. JAMA, 2020. 323(14): p. 1406–1407.

